# Reducing COVID-19 hospitalization risk through behavior change

**DOI:** 10.1101/2020.07.21.20159350

**Authors:** Mary L. Adams, David L. Katz, Joseph Grandpre, Douglas Shenson

## Abstract

Our objective was to determine strategies that could potentially reduce the risk of hospitalizations from COVID-19 due to underlying conditions. We used data (N=444,649) from the 2017 Behavioral Risk Factor Surveillance System to identify potentially modifiable risk factors associated with reporting any of the underlying conditions (cardiovascular disease, asthma, chronic obstructive pulmonary disease, diabetes, hypertension or obesity) found to increase risk of US hospitalizations for COVID-19. Risk factors included lifetime smoking, sedentary lifestyle, and inadequate fruit and vegetable consumption. Multiple logistic regression in Stata produced adjusted odds ratios (AORs) used to estimate population attributable-risk (PAR) in Excel. PARs for the 3 risk factors ranged from 12.4% for inactivity to 15.6% for diet for a combined PAR of 36.3%, implying that total elimination of these 3 risk factors could potentially reduce underlying conditions as much as 36%. This suggests that reducing COVID-19 hospitalizations might be a measurable and feasible US goal for the coronavirus pandemic. The simple lifestyle changes of increasing physical activity and fruit and vegetable consumption could reduce obesity, a key underlying condition and risk factor for 4 others. Reducing obesity and inactivity may also boost immunity. With uncertainly around how long the pandemic might last, other proposed strategies include wearing face masks when social distancing is not feasible, and addressing the special issues for nursing home residents. Such actions have the potential to lessen the impact of COVID-19 in the short term along with providing long term health benefits regarding chronic conditions.

## Introduction

One of the novel aspects of the current coronavirus pandemic is that information is constantly being updated as more studies are published. Based on early data from China^1^ people at risk of complications or death from COVID-19 include adults with cardiovascular disease (CVD), diabetes, chronic respiratory disease, asthma, hypertension, and cancer. Recent US data on hospitalizations for COVID-19 indicate a slightly different list of conditions affecting risk, with obesity added and cancer subtracted.^2^ Those results indicate that 89% of those hospitalized for COVID-19 had an underlying condition, the most common being hypertension, obesity, chronic lung disease (including asthma and chronic obstructive pulmonary disease), diabetes, and cardiovascular disease.^2^ Estimates of the percentage of adults who report any of those 6 underlying conditions are 56.0% overall, with ranges across states and demographic groups.^3^ The Guidelines for Opening up America Again released by the White House on April 16, 2020^4^ include a similar list to define vulnerable individuals, except that list includes elderly (undefined) and those with a compromised immune system and omits cardiovascular disease. And a recent study covering over a million US hospitalizations found that adults with a similar list of underlying conditions were 6 times as likely as those without underlying conditions to be hospitalized and 12 times as likely to die from COVID-19.^5^

Our objective for this study was to identify potentially modifiable risk factors that increase the likelihood of reporting any of the 6 underlying conditions (cardiovascular disease, asthma, chronic obstructive pulmonary disease, diabetes, hypertension or obesity) found to increase risk of US hospitalizations for COVID-19. ^2,3^ Preliminary unadjusted results show that older adults, men, and African-Americans were more likely to report an underlying condition, which are all groups that hospitalization data^2^ suggested were disproportionately affected by COVID-19. Each of the risk factors of lifetime smoking, sedentary lifestyle, and inadequate fruit and vegetable consumption in unadjusted results was associated with increased likelihood of reporting any of the 6 underlying conditions and an additive effect was suggested. ^3^ This study adds multiple logistic regression and population attributable-risk (PAR) to the analysis to estimate the proportion of the outcome that is caused by the risk factor or exposure. PARs take into account not only the relative risk of the factor but also the prevalence in the population and are useful in determining potential benefits of interventions as they suggest how much the outcome could be reduced by elimination or partial elimination of the factor.^6^ For example, Barnes and Yaffe estimated that 7% of Alzheimer’s disease cases in the US were potentially attributable to not completing high school.^7^ This study also adds measures of social determinants of health (SDOH): income < $25,000; not completing high school; African American or American Indian or Alaska Native; underinsurance; and living in the South or Midwest census region as opposed to the Northeast or West. Although causality has not been shown for each risk factor for each of the conditions included in our outcome, causal associations have been shown for all risk factors for at least one of the 6 outcomes included in our definition.^6,8^ We also assumed that all chosen SDOH measures had a role in causation for at least one of the separate outcomes. Depending on study results, our goal was to develop strategies to potentially reduce COVID-19 hospitalizations and deaths.

## Methods

### Data

We used publicly available 2017 Behavioral Risk Factor Surveillance System (BRFSS) data ^9^ from 444,649 adults ages 18 and older in the 50 states and DC. Data were adjusted for the probability of selection and weighted to be representative of the adult population in each state by age, gender, race/ethnicity, marital status, education, home ownership, and telephone type and included weights and stratum variables needed for analysis. Reliability and validity of the BRFSS have been found to be moderate to high for many survey measures.^10^ The median response rate for cell phone and land line surveys combined for the states included was 47.2%, ranging from 33.9% in California to 61.1% in Utah.^11^

### Measures

The outcome measure was used previously^3^ and includes adults reporting any of cardiovascular disease (CVD), asthma, chronic obstructive pulmonary disease (COPD), diabetes, hypertension, and obesity, with body mass index >30.0 kg/m^2^ based on self-reported height and weight. Risk factor measures were consistent with 3 of 4 upstream behavioral risk factors: ^6^ tobacco use, diet, and nutrition, and physical inactivity and their relationships^6,8^ to the components of our outcome are shown in Figure 1. Ever smoking included respondents who smoked 100 cigarettes in their lifetime whether they currently smoked or not. Inadequate fruit and vegetable consumption was defined as consuming the combination <5 times per day based on responses to five separate questions and respondents who did not participate in any leisure time physical activity in the past month were considered to have a sedentary lifestyle. Once unknowns were removed, final N’s for the 3 separate risk factors ranged from 389,200 for fruit and vegetable consumption to 426,403 for reporting ever smoking.

**Figure 1.**
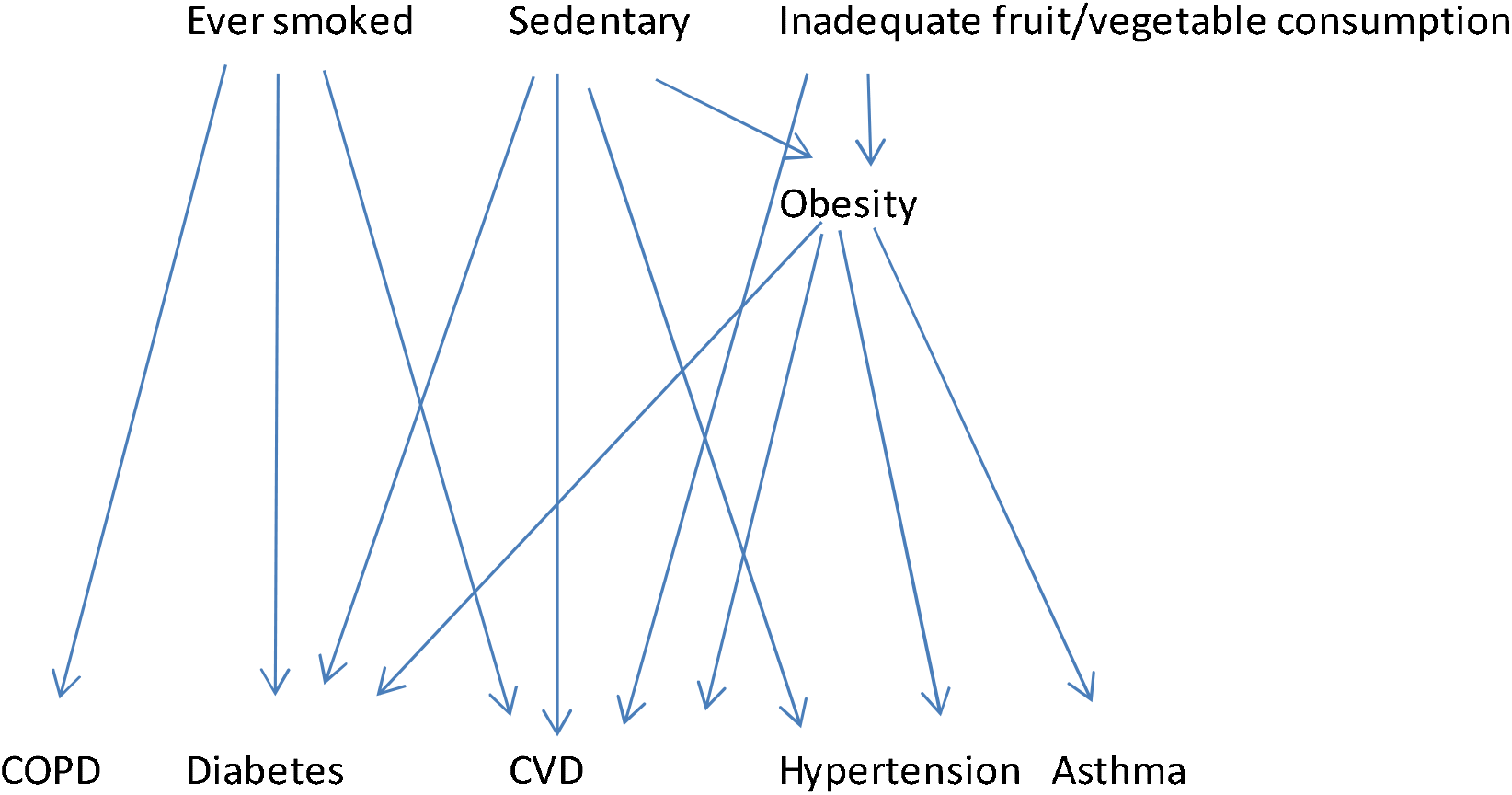
Diagram showing relationship between risk factors and underlying conditions Abbreviations: CVD: cardiovascular disease; COPD: chronic obstructive pulmonary disease

Our SDOH measures were consistent with the 5 general categories in Healthy People 2020^12^ and limited by availability on the BRFSS. SDOH measures were 1. household income < $25,000 vs all else including unknown; 2. not completing high school; 3. African American or American Indian or Alaska Native vs. all else; 4. underinsurance, defined as either no health insurance or needing to see a doctor in the past year but unable to, due to cost; and 5. living in the South or Midwest census region as opposed to the Northeast or West. Measures 3 and 5 were defined based on preliminary analysis^3^ of the data showing these groups at higher risk for reporting any of the underlying conditions.

Demographic measures included gender and age (18-24, 25--34, 35-44, 45-54, 55-64, 65-74, and 75 years and older).

### Analysis

Stata version 14.1 (Stata Corp LP, College Station, TX) was used for data analysis to account for the complex sample design of the BRFSS. Point estimates and 95% confidence intervals are reported for cross tabulations for the composite measure and each component outcome by each of the SDOH measures and risk factors. Logistic regression was done to obtain adjusted odds ratios (AORs) to use in estimating population-attributable risk (PAR) using Levin’s formula^13^ and Excel. Refusals and “don’t know” responses are removed unless otherwise indicated (income). Composite measures of the 3 risk factors and the 5 SDOH measures were created to determine if a linear response was confirmed by multiple logistic regression. A combined PAR was computed using the following formula to avoid totals >100%:^7,14^

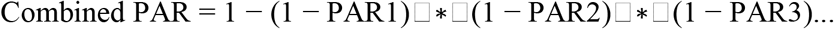

## Results

Overall prevalence of reporting any of the underlying conditions was 56.0% (95% CI 55.7–56.3) with results for demographic groups available elsewhere.^3^ Prevalence rates of the separate components were 8.5% for CVD, 6.6% for COPD, 9.1% for asthma, 10.8% for diabetes, 32.4% for hypertension, and 30.1% for obesity, while for risk factors were 40.4% for ever smoking, 26.6% for sedentary lifestyle, 84.1% for inadequate fruit and vegetable consumption; 91.1% of study adults reported ≥1 of the 3 risk factors. Substituting current smoking with a prevalence of 16.4%, 88.1%, of adults reported ≥1 of the 3 risk factors. For the SDOH measures, 13.5% did not finish high school, 23.2% reported income <$25,000, 20.9% were underinsured, 58.7% lived in the South or Midwest, 13.0% were Black or American Indian and 75.9% reported ≥1. Unadjusted results (Table 1) show that all 3 risk factors were directly associated with the key outcome and with all its components except asthma, where consumption of fruits and vegetables was not significant. Among SDOH measures, income, education, and race were directly associated with all outcomes, while being underinsured was protective or had no effect for the key outcome and 3 of its components, and living in the South or Midwest increased risk for all outcomes except asthma, where region was slightly but significantly protective.

**Table 1.** Unadjusted results for risk factors social determinants of health (SDOH) measures and selected outcomes 2017 Behavioral Risk Factor Surveillance System; 50 states; N>400,000

| Risk factor | Any underlying |  | CVD |  | Diabetes |  | COPD |  | Obese |  | Asthma |  | HBP |  |
| --- | --- | --- | --- | --- | --- | --- | --- | --- | --- | --- | --- | --- | --- | --- |
|  | % | CI | % | CI | % | CI | % | CI | % | CI | % | CI | % | CI |
| Ever smoked | 64.3 | 63.8-64.8 | 12.8 | 12.5-13.1 | 13.3 | 12.9-13.6 | 12.4 | 12.1-12.7 | 32.3 | 31.8-32.8 | 10.7 | 10.4-11.0 | 39.4 | 38.9-39.9 |
| Never smoked | 50.5 | 50.0-51.0 | 5.7 | 5.5-5.9 | 9.2 | 9.0-9.4 | 2.8 | 2.7-3.0 | 28.7 | 28.3-29.1 | 8.0 | 7.8-8.3 | 28.0 | 27.6-28.4 |
| Sedentary | 67.8 | 67.2-68.5 | 12.7 | 12.3-13.1 | 16.2 | 15.7-16.7 | 11.4 | 11.0-11.7 | 39.6 | 38.9-40.2 | 11.1 | 10.8-11.5 | 41.3 | 40.7-42.0 |
| Not sedentary | 52.3 | 51.9-52.7 | 7.1 | 6.9-7.3 | 9.0 | 8.8-9.2 | 5.0 | 4.9-5.2 | 27.0 | 26.6-27.4 | 8.4 | 8.2-8.6 | 29.6 | 29.3-30.0 |
| Eat < 5 | 57.0 | 56.6-57.4 | 8.6 | 8.5-8.8 | 11.1 | 10.9-11.4 | 6.9 | 6.7-7.0 | 31.1 | 30.7-31.4 | 9.0 | 8.8-9.2 | 33.3 | 33.0-33.7 |
| Eat 5 or more | 50.0 | 49.0-50.9 | 6.9 | 6.6-7.3 | 8.7 | 8.3-9.3 | 5.0 | 4.6-5.3 | 25.8 | 25.0-26.6 | 9.4 | 8.9-9.9 | 27.7 | 26.9-28.4 |

| SDOH factor | Any underlying |  | CVD |  | Diabetes |  | COPD |  | Obese |  | Asthma |  | HBP |  |
| --- | --- | --- | --- | --- | --- | --- | --- | --- | --- | --- | --- | --- | --- | --- |
|  | % | CI | % | CI | % | CI | % | CI | % | CI | % | CI | % | CI |
| <High school | 66.8 | 65.6-67.9 | 13.5 | 12.9-14.2 | 17.2 | 16.5-18.0 | 11.3 | 10.7-11.8 | 35.6 | 34.5-36.8 | 11.0 | 10.4-11.6 | 39.1 | 38.1-40.2 |
| High school grad | 54.5 | 54.1-54.9 | 7.8 | 7.6-7.9 | 9.8 | 9.6-10.0 | 5.8 | 5.7-6.0 | 29.3 | 29.0-29.6 | 8.8 | 8.6-9.0 | 31.4 | 31.1-31.7 |
| Income<\$25,000 | 65.1 | 64.4-65.9 | 13.2 | 12.8-13.7 | 15.8 | 15.3-16.3 | 11.7 | 11.3-12.1 | 35.2 | 34.5-35.9 | 12.2 | 11.8-12.7 | 38.4 | 37.7-39.0 |
| Income \$25,000+ | 53.3 | 52.9-53.7 | 7.1 | 7.0-7.3 | 9.4 | 9.2-9.6 | 5.1 | 4.9-5.2 | 28.6 | 28.2-28.9 | 8.1 | 7.9-8.3 | 30.7 | 30.4-31.0 |
| Underinsured | 56.4 | 55.6-57.3 | 8.0 | 7.6-8.4 | 9.7 | 9.3-10.2 | 7.9 | 7.5-8.2 | 33.1 | 32.3-33.8 | 10.5 | 10.1-11.0 | 28.0 | 27.4-28.7 |
| Not underinsured | 56.0 | 55.6-56.4 | 8.7 | 8.5-8.8 | 11.1 | 10.9-11.3 | 6.2 | 6.1-6.4 | 29.4 | 29.1-29.7 | 8.7 | 8.5-8.9 | 33.6 | 33.3-34.0 |
| South/Midwest | 58.4 | 57.9-58.8 | 9.3 | 9.1-9.5 | 11.4 | 11.1-11.6 | 7.4 | 7.2-7.6 | 32.4 | 32.0-32.8 | 8.9 | 8.6-9.1 | 34.3 | 33.9-34.7 |
| Northeast/West | 52.6 | 52.1-53.2 | 7.4 | 7.2-7.6 | 10.0 | 9.7-10.4 | 5.4 | 5.2-5.6 | 26.8 | 26.3-27.3 | 9.3 | 9.0-9.6 | 29.7 | 29.2-30.2 |
| AA_AI/AN | 65.9 | 64.9-66.9 | 9.3 | 9.1-9.5 | 14.6 | 13.9-15.2 | 7.0 | 6.6-7.5 | 39.0 | 38.0-39.9 | 11.8 | 11.2-12.5 | 40.9 | 40.0-41.8 |
| Other | 54.5 | 54.1-54.9 | 7.4 | 7.2-7.6 | 10.3 | 10.1-10.5 | 6.5 | 6.3-6.6 | 28.8 | 28.5-29.1 | 8.6 | 8.5-8.8 | 31.1 | 30.8-31.4 |

Not positively associated with outcome
Abbreviations: CVD: cardiovascular disease; COPD: chronic obstructive pulmonary disease; HBP: high blood pressure; SDOH: social determinants of health.

Results of multiple logistic regression (Table 2) for any underlying condition indicate that the highest adjusted odds ratio (AOR) was 7.3 (6.8-7.8) for adults ages 70-79 and all measures included in the model had AORs of 1.12 or higher. Thus adjusted results confirm all unadjusted results except for underinsurance which is significantly associated with higher risk in adjusted results. Adjusted results also add men to those at significantly higher likelihood of reporting an underlying condition. If a composite measure is used in the models instead of the separate risk factor measures, the AOR for all 3 risk factors vs. none is 2.62 (2.43-2.82) and that for all 5 SDOH measures is 2.51 (1.68-3.73). Logistic regression results for the 6 underlying conditions are available in supplemental tables. Results for PARs for the 3 risk factors for any underlying condition plus each component condition are shown in Table 3 indicating combined PARs ranging from 20.4% for asthma to 66.9% for COPD. At least 2 of the 3 risk factors contribute attributable-risk for each outcome and all 3 contribute a combined 36.3% for any of the 6. Combined PARs for SDOH measures ranged from 13.5% for asthma to 37.5% for COPD and 29.2% for any of the 6.

**Table 2.** Adjusted odds ratios (AOR) from multiple logistic regression with all measures included in the model with outcome any of 6 underlying conditions ^a^.

N=346,198.
| Group | Odds Ratio | 95% CI |
| --- | --- | --- |
| Females (referent) |  |  |
| Males | 1.15 | 1.11-1.19 |
| Age 18-29 (referent) |  |  |
| 30-39 | 1.58 | 1.49-1.67 |
| 40-49 | 2.21 | 2.09-2.34 |
| 50-59 | 3.31 | 3.13-3.50 |
| 60-69 | 4.85 | 4.59-5.13 |
| 70-79 | 7.30 | 6.84-7.80 |
| 80+ | 7.19 | 6.51-7.95 |
| High school grad (Referent) |  |  |
| < High school | 1.26 | 1.18-1.35 |
| Income ≥\$25,000 (Referent) | | |
| Income <\$25,000 | 1.46 | 1.40-1.53 |
| Not underinsured (Referent) |  |  |
| Underinsured | 1.12 | 1.07-1.17 |
| White, Hispanic, Asian, Other (Referent) |  |  |
| Black or American Indian | 1.70 | 1.61-1.80 |
| West or Northeast (Referent) |  |  |
| South or Midwest | 1.18 | 1.14-1.21 |
| Never smoked Referent) |  |  |
| Ever smoked | 1.40 | 1.35-1.45 |
| Not sedentary (Referent) |  |  |
| Sedentary | 1.53 | 1.47-1.60 |
| Eat F/V 5X a day (Referent) |  |  |
| Eat <5 a day | 1.22 | 1.16-1.27 |

<sup>a</sup> Cardiovascular disease, diabetes, chronic obstructive pulmonary disease, asthma, hypertension, and/or obesity.

**Table 3.** Population attributable risk (PAR) for the outcome of any underlying condition ^a^ and its components; 2017 Behavioral Risk Factor Surveillance System; N=346,198

| Measure | PAR |  |  |  |
| --- | --- | --- | --- | --- |
|  | Ever smoked | Sedentary | Eat<5 | Combined <sup>b</sup> |
| Any underlying condition | 13.9% | 12.4% | 15.6% | 36.3% |
| Cardiovascular disease | 24.0% | 8.7% | 0.0% | 30.6% |
| Hypertension | 9.2% | 8.3% | 13.1% | 27.7% |
| Diabetes | 2.8% | 11.5% | 8.5% | 21.2% |
| Obesity | 1.6% | 14.5% | 14.4% | 28.0% |
| COPD | 55.8% | 16.3% | 10.5% | 66.9% |
| Asthma | 15.1% | 6.2% | 0.0% | 20.4% |

Abbreviations: PAR: Population attributable- risk; COPD: chronic obstructive pulmonary disease; Eat<5: Eat fruits and vegetables < 5 times/day.
<sup>a</sup> Cardiovascular disease, diabetes, chronic obstructive pulmonary disease, asthma, hypertension, and/or obesity.
<sup>b</sup> Combined PAR

## Discussion

The list of underlying conditions has changed over time but since using US hospitalization data which added obesity ^2,3^ to the original list based on data from China^1^ the major conditions on the list ^4,5^ remain fairly constant as the ones chosen for study. In at least one large and recent study, hypertension was included in cardiovascular disease rather than listed as a separate condition.^5^ Excluding age as a criterion for risk allows the study of age associations. In our adjusted results, older age, being male, African American, American Indian, not finishing high school, having income <$25,000, being underinsured, and living in the South or Midwest census regions are all significantly associated with reporting any of the 6 underlying conditions. The first three listed are consistent with US data showing that older adults, men, and African Americans are disproportionately among those hospitalized for COVID-19 and suggest that the reason may be due to their being more likely to report an underlying condition.

PAR results for the 3 potentially modifiable risk factors are consistent with results for the actual causes of deaths which have been reported as tobacco, diet and physical activity.^15^ Figure 1 illustrates the relationship between the 3 upstream risk factors and the underlying conditions, with obesity as an intermediate outcome for 4 of the 6. Other outcomes such as cognitive decline and dementia have also been shown to be associated with these risk factors.^7,8,16^ Realistically, changing behavior is going to have limited effectiveness in the short term but in the long term may reduce risk and thus hospitalizations and deaths from COVID-19 and other chronic conditions shown to be associated with these risk factors.^6-8^ Also it is important to realize that risk factor reduction will be most effective for the 44% of adults who do not already have an underlying condition and that only current smoking (not lifetime) is potentially modifiable. However, a significant short term effect is possible from simply not being totally sedentary and adding more fruits and vegetables to the diet to reduce obesity, which is one of the most prevalent of the underlying conditions increasing risk of hospitalizations.^2^ In addition, as noted in Figure 1, obesity is a risk factor for CVD, asthma, diabetes, and hypertension. As shown in Table 3, total elimination of all 3 obesity risk factors would reduce obesity by 28%; not shown is that if everyone did some exercise and ate fruits and vegetables 5 times each day, obesity could be reduced by 26.9% to below 22%. If HALF of all adults made those changes, obesity prevalence could be reduced below 26% with the potential for reductions in the 4 other underlying conditions for which obesity is a risk factor.

Results from other studies suggest additional benefits from increasing exercise and fruit and vegetable consumption. A study of influenza epidemics found that being overweight increases risk of infection, so concluded that losing weight and engaging in moderate exercise can boost immunity.^17^ Our results suggest that increasing fruit and vegetable consumption can help weight loss efforts. Quitting smoking may reduce any direct effects of smoking on COVID-19 complications, as ICU admissions have been reported to be more common among smokers.^18^ Strategies to reduce these risk factors such as public health campaigns could be done within current pandemic guidelines through ad campaigns using various media outlets. It seems that most conversations about COVID-19 have focused on stopping its spread or treating it symptoms with little or no attention to how major complications might be prevented.

Estimated PARs for the SDOH measures are lower than those for risk factors and potentially not as easy to modify especially in the short term. But they serve as a reminder that the risk factor measures were controlled for age, gender and the 5 SDOH measures and vice versa. So for example, higher rates of underlying conditions in the South and Midwest compared with the West and Northeast were controlled for any differences in risk factors, education, income, etc. Further study will be needed to determine if American Indians, lower income adults, those without a high school diploma, the underinsured, and residents of Southern and Midwestern states also have higher *hospitalization* rates compared with those not in those groups. Being underinsured could create separate issues with access to care and treatment options especially since these underlying conditions are likely considered pre-existing conditions by most healthcare insurers.

In addition of the potential of lowering risk through lifestyle change, there is evidence that among hospitalized adults with COVID-19 and diabetes^19^ those with well-controlled blood glucose had lower risk of undesirable outcomes compared with less well-controlled levels. Among hospitalized patients with COVID-19 and coexisting hypertension, use of ACEI/ARB was associated with lower risk of all-cause mortality compared with ACEI/ARB nonusers.^20^ Thus ongoing management of existing underlying conditions to reduce risk as much as possible should be an objective. Quantitating any reduction in hospitalizations due to risk factor reduction or better management of diabetes or hypertension will be extremely difficult, especially while the pandemic continues.

Many uncertainties remain with the coronavirus including when and how the pandemic will end. Perhaps an effective vaccine will be found that enough US adults will accept or a treatment will be discovered that greatly reduces the impact of COVID-19. But it is probably safe to assume it will not disappear on its own anytime soon and meanwhile life must go on. Results of this study suggest merit in focusing on the more than half of adults with underlying conditions that increase their risk of hospitalizations and defining a goal for the US coronavirus pandemic as reducing hospitalizations and deaths. As results from this study suggest, there are a variety of potential options to achieve that goal. We suggest that a focus on risk factors for underlying conditions is warranted as shown by results and cited studies in this paper. Any possible reduction in COVID-19 *cases* due to a possible boost in immunity would be seen as a bonus.^17^ In addition to addressing the 3 risk factors and management of diabetes and hypertension, general strategies to reduce the spread of the disease should be continued or strengthened where needed. These include social distancing and wearing masks^21^ which have been shown to be quite effective. About 49%^3^ of employed adults have an underlying condition so some accommodation may be needed to protect these employed adults from acquiring COVID-19 or the consequences of losing their income if they chose not to return to work. Because a large fraction of COVID-19 deaths in the US have been among the 1.3 million patients in nursing homes^22^ plans for this population will be critical and are already being proposed.^23^ We hope that they will include reporting more COVID-19 data (e.g. hospitalizations) separately for nursing home patients as they currently do for deaths. We also suggest that consideration be given to vaccinating all nursing home patients with MMR vaccine. Studies and observations have suggested that the MMR vaccine might offer some protection against COVID-19 or at least its most severe consequences.^24,25^

### Limitations

There are several limitations to this study including variability in reliability and validity of the self-reported data.^10^ Validity was usually high when compared to medical records but obesity and current tobacco use showed some differences between self-reports and physical measures.^10^ It is possible that additional conditions will be found to be associated with COVID-19 hospitalizations and death, some of which might not be available on the BRFSS. PAR estimates assume there is a causal relationship and this may not be true for the SDOH measures. Persons who ever smoked were grouped together with no differentiation for the length of time smoked or how long it might have been since they quit which may result in imprecision in the accuracy of the attributable-risk estimates for smoking. The generalizability of these results is unknown but there is no reason to believe it would not be good. Another study limitation is that only non-institutionalized adults were surveyed so the 1.3 million adults ^23^ in nursing homes who may be more likely to have the outcomes studied were excluded. Household adults who are physically or mentally unable to respond to a survey are also excluded which may omit some potential respondents with these underlying conditions.^26^ The effect of excluding persons unable to respond to a telephone survey implies that adults in this current study may be less affected by their outcomes than other adults in the community.

## Conclusion

Based on evidence from this and other studies, we propose a measureable US goal for addressing the coronavirus pandemic of reducing hospitalizations and death by focusing on 3 risk factors for the 6 underlying conditions found to be associated with increased risk of hospitalizaions.^2^ Short-term strategies for non-smokers to increase physical activity and fruit and vegetable consumption could perhaps even boost immunity and reduce the number of COVID-19 cases while reducing obesity. Attention should also be given to improving blood glucose control in adults with diabetes and blood pressure control among those with hypertension. These results suggest that along with the wearing of face masks and/or physical distancing to help prevent the spread of COVID-19, there are simple lifestyle changes that have the potential to lessen its impact.

## Data Availability

Data and documentation are available at this link: https://www.cdc.gov/brfss/annual_data/annual_2017.html

## Acknowledgments

No funding was received for this study.

## References

1. Wu Z, McGoogan JM. Characteristics of and important lessons from the coronavirus disease 2019 (COVID-19) outbreak in China: summary of a report of 72LJ314 cases from the Chinese Center for Disease Control and Prevention. JAMA. Published February 24, 2020. doi:10.1001/jama.2020.2648

2. Garg S, Kim L, Whitaker M, O’Halloran A, Cummings C, Holstein R, et al. Hospitalization rates and characteristics of patients hospitalized with laboratory-confirmed coronavirus disease 2019—COVID-NET, 14 States, March 1–30, 2020. MMWR Morb Mortal Wkly Rep. 2020;69:458–64. PubMed https://doi.org/10.15585/mmwr.mm6915e3

3. Adams ML, Katz DL, Grandpre J. Updated estimates of chronic conditions affecting risk for complications from coronavirus disease, United States. Emerg Infect Dis. 2020 Sep [date cited]. https://doi.org/10.3201/eid2609.202117

4. The White House (Washington, DC). Guidelines Opening Up America Again. April 16, 2020. Available at: https://www.whitehouse.gov/openingamerica/?fbclid=IwAR2m39LPMgNUAcDLvjRR3DHLh1Lhwxlc-o3BN3Eous0FSeagdsuj2Jwuf3U Accessed April 18, 2020.

5. Stokes EK, Zambrano LD, Anderson KN, et al. Coronavirus Disease 2019 Case Surveillance — United States, January 22–May 30, 2020. MMWR Morb Mortal Wkly Rep. ePub: 15 June 2020. DOI: http://dx.doi.org/10.15585/mmwr.mm6924e2

6. Brownson RC, Remington PL, Wegner, MV. Chronic Disease Epidemiology and Control. 4th ed. Washington, DC: American Public Health Association; 2016.

7. Barnes DE, Yaffe K. The projected effect of risk factor reduction on Alzheimer’s disease prevalence. Lancet Neurol. 2011 Sep;10(9):819–28. doi:10.1016/S1474-4422(11)70072-2.

8. Adams M, Grandpre J, Katz D, Shenson D. The impact of key modifiable risk factors on leading chronic conditions. Prev Med. 2019 Mar;120:113–118. https://doi.org/10.1016/j.ypmed.2019.01.006.

9. Behavioral Risk Factor Surveillance System (BRFSS) (Atlanta, Georgia). Centers for Disease Control and Prevention. 2017 Survey Data and Documentation. https://www.cdc.gov/brfss/annual_data/annual_2017.html. Accessed November 16, 2018.

10. Pierannunzi C, Hu SS, Balluz L. A systematic review of publications assessing reliability and validity of the Behavioral Risk Factor Surveillance System (BRFSS), 2004-2011. BMC Med Res Methodol. 2013 Mar 24;13:49. Doi: 10.1186/1471-2288-13-49.

11. Behavioral Risk Factor Surveillance System (BRFSS) (Atlanta, Georgia). Centers for Disease Control and Prevention. Behavioral Risk Factor Surveillance System 2017 Summary Data Quality Report, June 13, 2018. https://www.cdc.gov/brfss/annual_data/2017/pdf/2017-sdqr-508.pdf, accessed November 16, 2018.

12. U.S. Department of Health and Human Services (Washington, DC): Healthy People 2020.Social Determinants of Health. https://www.healthypeople.gov/2020/topics-objectives/topic/social-determinants-of-health. Accessed May 27, 2019.

13. Rückinger S, von Kries R and Toschke AM. An illustration of and programs estimating attributable fractions in large scale surveys considering multiple risk factors. BMC Med Res Methodol. 2009; 9: 7. doi:10.1186/1471-2288-9-7.

14. Rowe AK, Powell KE, Flanders WD. Why population attributable fractions can sum to more than one. Am J Prev Med. 2004 Apr;26(3):243–9.

15. Mokdad AH, Marks JS, Stroup DF, Gerberding JL. Actual causes of death in the United States, 2000. JAMA. 2004 Mar 10;291(10):1238–45. Correction in JAMA. 2005 Jan 19;293(3):298.

16. Adams ML, Grandpre J, Katz DL, Shenson D. Cognitive Impairment and Cardiovascular Disease: A Comparison of Risk Factors, Disability, Quality of Life, and Access to Health Care. Public Health Rep. 2020 Jan;135(1):132–140. doi:10.1177/0033354919893030.

17. Luzi L, Radaelli MG. Influenza and Obesity: Its Odd Relationship and the Lessons for COVID-19 Pandemic. Acta Diabetol. 2020 Jun;57(6):759–764. doi:10.1007/s00592-020-301522-8. Epub 2020 Apr 5.

18. Engin AB, Engin ED and Engin A. Two important controversial risk factors in SARS-CoV-2 infection: Obesity and smoking. Environ Toxicol Pharmacol. 2020 Aug; 78: 103411. Published online 2020 May 15. doi:10.1016/j.etap.2020.103411

19. Zhu L, She ZG, Cheng X, Qin JJ, Zhang XJ, Cai J, Lei F. et al. Association of blood glucose control and outcomes in patients with COVID-19 and pre-existing type 2 Diabetes. Cell Metab. 2020 doi:10.1016/j.cmet.2020.04.021.

20. Zhang P, Zhu L, Cai J, Lei F, Qin JJ, Xie J, Liu YM. et al. Association of inpatient use of angiotensin converting enzyme inhibitors and angiotensin II receptor blockers with mortality among patients with hypertension hospitalized with COVID-19. Circ Res. 2020.

21. Chu DK, Akl EA, Duda S, Solo K, Yaacoub S, Schünemann HJ; COVID-19 Systematic Urgent Review Group Effort (SURGE) study authors. Physical distancing, face masks, and eye protection to prevent person-to-person transmission of SARS-CoV-2 and COVID-19: a systematic review and meta-analysis. Lancet. 2020 Jun 27;395(10242):1973–1987. doi: 10.1016/S0140-6736(20)31142-9. Epub 2020 Jun 1.

22. Centers for Disease Control and Prevention (Atlanta, GA). National Center for Health Statistics; Nursing Home Care. https://www.cdc.gov/nchs/fastats/nursing-home-care.htm.

23. American Geriatrics Society. American Geriatrics Society (AGS) Policy Brief: COVID-19 and Assisted Living Facilities. J Am Geriatr Soc. 2020 Jun;68(6):1131–1135 doi:10.1111/jgs.16510. Epub 2020 May 14.

24. Gold, Jeffrey. (2020). MMR Vaccine Appears to Confer Strong Protection from COVID-19: Few Deaths from SARS-CoV-2 in Highly Vaccinated Populations. 10.13140/RG.2.2.32128.25607.

25. Young A, Neumann B, Mendez RF, Reyahi A,et al. Homologous protein domains in SARS-CoV-2 and measles, mumps and rubella viruses: preliminary evidence that MMR vaccine might provide protection against COVID-19. Available at: https://www.medrxiv.org/content/10.1101/2020.04.10.20053207v1.full.pdf

26. Adams M. Results and their implications from comparing respondents and proxy responses for non-respondents with cognitive difficulties on a telephone survey. Disabil Health J. 2017 Jan;10(1):131–138. doi:10.1016/j.dhjo.2016.09.004. Epub 2016 Sep 13.

